# A Mendelian randomisation study of smoking causality in IPF compared with COPD

**DOI:** 10.1101/2020.12.04.20243790

**Authors:** Anna Duckworth, Michael A. Gibbons, Robin N Beaumont, Andrew R Wood, Howard Almond, Katie Lunnon, Mark A. Lindsay, Chris J Scotton, Jess Tyrrell

**Author notes:** These authors contributed equally (Chris J Scotton, Jess Tyrrell).

## Abstract

In a normal year, the fatal lung disease Idiopathic Pulmonary Fibrosis (IPF) accounts for ∼1% of UK deaths. Smoking is a recognised risk factor for IPF but the question of causality remains unanswered. Here, we used data from the UK Biobank (UKBB) and the well-established genetic technique of Mendelian randomisation (MR) methods to investigate whether smoking is causal for IPF compared with COPD, where causality is established.

We looked at observational associations in unrelated Europeans, with 871 IPF cases, 11,413 COPD cases and 366,942 controls. We performed analyses using one-sample MR to test for inferred smoking causality in ever smokers using genetic variants that have a previously demonstrated association with smoking heaviness.

Strong associations between disease status and ever having smoked were found in both IPF (OR = 1.52; 95%CI:1.32-1.74; P=2.4×10^−8^) and COPD (OR= 5.77; 95%CI:5.48-6.07; P<1×10^−15^). Using MR, a one allele increase in smoking volume genetic risk score was associated with higher odds of COPD in ever smokers, (OR = 4.32; 95%CI:3.37-5.54; P<1×10^−15^), but no association was seen in IPF (OR=0.55; 95%CI: 0.17-1.81; P=0.33). No association was found between the genetic risk score and disease prevalence in never smokers with IPF (OR = 1.00; 95%CI:0.98-1.02; P=1.00) or COPD (OR = 1.00; 95%CI:0.99-1.01; P=0.53).

Although both IPF and COPD are observationally associated with smoking, our analysis provides evidence inferring that the association is causal in COPD but there is no such evidence in IPF. This suggests that other environmental exposures also need consideration in IPF.

## Introduction

Idiopathic pulmonary fibrosis (IPF) is a fibrotic lung disease of exclusion; when a cause (such as asbestos) is identified, it is no longer classified as IPF. Smoking is cited as one of the key risk factors, along with age, being male [1, 2] and other environmental contributors (e.g. airborne dusts) [3] and yet the unique contribution of smoking to disease causality is not known. Chronic obstructive pulmonary disease (COPD) is another age-related respiratory disease which is firmly associated with smoking or exposure to burning biomass [4], with additional evidence suggestive of a link with air pollution [5].

In COPD, the strong causal link with smoking derives from evidence that the most effective treatment is smoking cessation [4]. In contrast, the impact of smoking on IPF prognosis has conflicting reports. In 2001, King et al showed differing median survivals of 116.4, 25.3 and 27.2 months for current, former and never smokers respectively (N=238, p=0.0003) [6] – although smokers were significantly younger at diagnosis. A 2016 study (N=98) reported that never-smoking IPF patients developed more acute exacerbations than smokers (50% vs 18.2%, p<0.0001), with median survival times of 18.5 and 26.3 months respectively, P<0.0001 (with similar diagnosis age) [7]. This contrasts with a 2008 study (N=249), in which survival was evaluated against smoking status using proportional hazards analysis, adjusting for sex, age, disease severity and degree of honeycombing. Although current smokers had milder disease on presentation than former smokers, with associated higher unadjusted survival, HR=0.44 [90% CI:0.24-0.80, P=0.007], after adjustment for the morphologic extent of fibrosis, survival was the same for former and current smokers and higher in never smokers [8]. Two recent large studies (N=453, N=1263) reported no impact on survival [9, 10].

As the above results illustrate, deducing a causal relationship from retrospective observational data is difficult and can be complicated by confounding; prospective studies to establish causality with smoking exposure are clearly not feasible. As an alternative, Mendelian randomisation (MR) has become a well-established approach for examining causal inferences in large data sets, using genetic variants as proxies for the exposure [11]. We chose to investigate the question of smoking causality in IPF and COPD using MR with UK Biobank (UKBB) data. Analysis of smoking causality using MR has previously been reported for COPD in the context of a schizophrenia study [12] and for systemic inflammation and airway limitation [13].

## Material and methods

### Collection and selection of UKBB Data

The UK Biobank is a repository of research data sourced from ∼500,000 UK-wide volunteers (99.5% aged 40-69), recruited during 2006-2010 [14]. Participants provided data during registration via questionnaire, interview, physical measurements and samples for biomarker and genetic analysis, with long-term follow-up. Patient and public involvement information is available online [15]. Hospital record data were linked to 31/3/2017. To avoid genetic biases, we restricted our analysis to a subset of 379,708 unrelated individuals of white European descent. Genotyping and cohort derivation is described in Supplement S1. We defined IPF cases as having a primary or secondary ICD10 code HES (Hospital Episodes Statistics) diagnosis of J84.0 (Alveolar and parieto-alveolar conditions), J84.1 (Other interstitial pulmonary disease with fibrosis), J84.8 (Other specified interstitial pulmonary diseases) and J84.9 (Interstitial pulmonary disease, unspecified): 1,353 unrelated cases in total. We defined COPD cases as having a primary or secondary ICD10 code of J41 (Simple and mucopurulent chronic bronchitis), J42 (Unspecified chronic bronchitis), J43 (Emphysema) or J44 (Other chronic obstructive pulmonary disease) plus those self-reported to have COPD: a total of 11,895 unrelated cases. We excluded 482 cases with both IPF and COPD, to leave 11,413 COPD cases and 871 IPF cases. Controls were defined as participants without COPD or IPF (n=366,942). Survival data was derived from linked death registry statistics (also to 31/3/17).

### Observational Associations

Derivation of all variables is decribed in Supplement S1. Full details of the spirometry protocol are available [16]. Lung function percent predicted values were calculated using Global Lung Initiative equations [17]. All data values were recorded at UKBB registration and not at diagnosis.

We used logistic regression models in Stata 16.0 to compare the key demographics of the IPF and COPD groups with controls (adjusting for age and sex throughout) and to investigate the association with smoking volume (in pack years, transformed to log_10_pack years for linear regression). Mortality was calculated with death as the outcome variable. Smoking history is reported in 10-pack-year bins. The continuous measure of smoking used in the MR analyses, log pack years, was adjusted for age, sex, principal components, assessment centre and the residuals were then taken and inverse normalised.

In order to explore indirect smoking associations with IPF, we also investigated parental smoking. This is only specifically recorded in UKBB as maternal smoking during pregnancy so we analysed the association of IPF prevalence with this and also with paternal and maternal lung cancer incidence (acknowledging that not all will be a consequence of smoking), both using logistic regression adjusted for age and sex. We also determined whether maternal smoking during pregnancy and paternal lung cancer prevalence are associated with smoking in offspring.

### Mendelian Randomisation

MR [18] can be used to test for a causal relationship from a behaviour that could be genetically influenced, such as smoking amount, to a disease outcome, such as IPF or COPD. Causality can be inferred in one direction because genetic make-up is randomly allocated at conception and unlikely to be influenced by environmental factors, such as diet and pollution. Figure 1 illustrates the MR principle and its application here.

**Figure 1.**
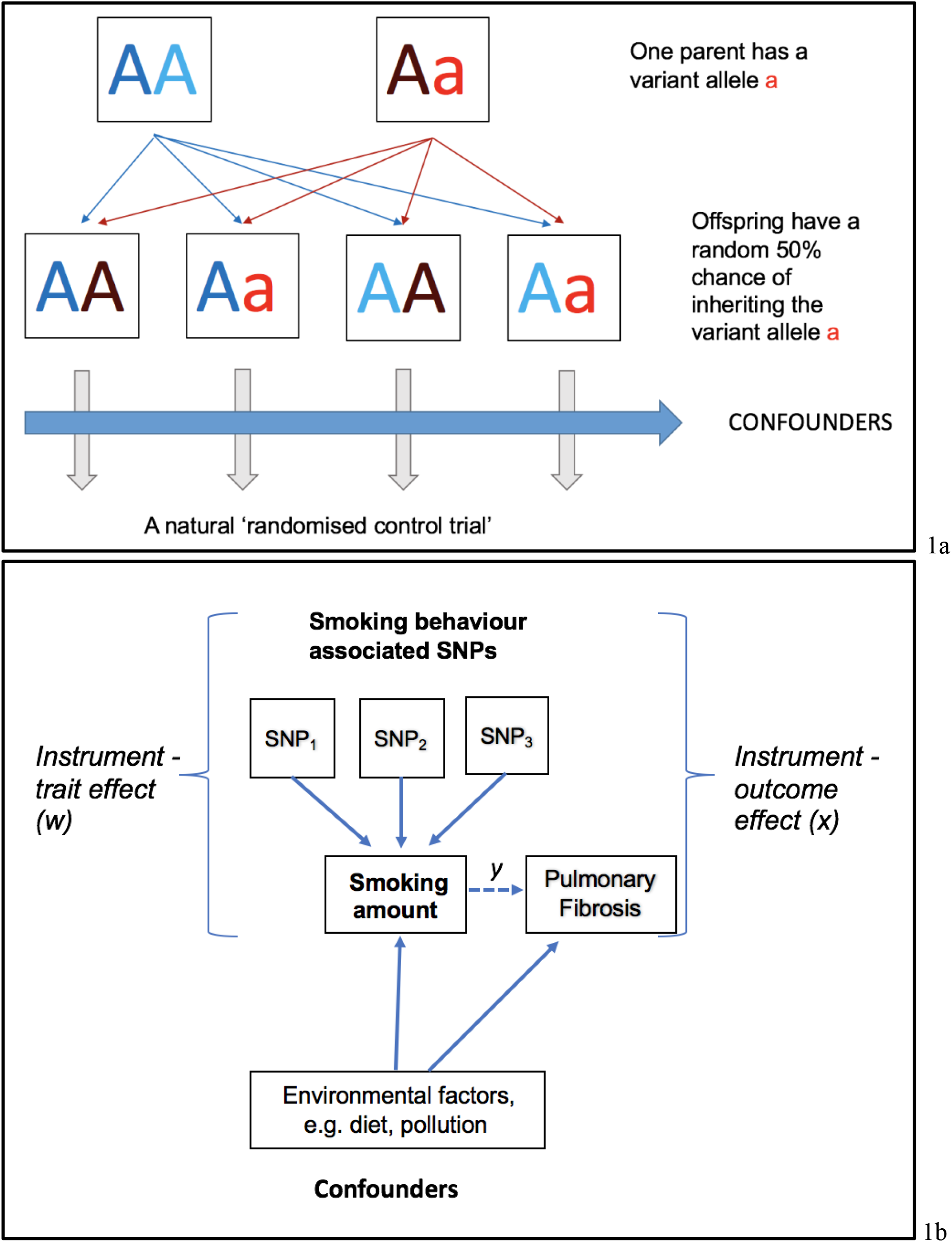
Principle of Mendelian Randomisation. (a) Illustration demonstrating how Mendelian inheritance leads to a random allocation of variant alleles amongst offspring such that confounders affecting different social groups across the whole sample can be controlled for statistically; akin to a natural randomised control trial (b) If a trait such as smoking intensity (X) plays a causal role in the disease outcome IPF or COPD (Y), genetic variants (Z_i_) associated with smoking intensity will also be associated with the outcome Y. Since genotype is assigned at conception, it should not be associated with environmental risk factors that would normally confound the association between the trait and lung disease (eg diet or pollution). Weighted estimates of the genetic-trait association (w) and the genetic-lung disease association (x) can be used to infer the causal effect of that trait on the lung disease (y=x/w), which is expected to be free from confounding.

### Identification of genetic instrument variants

Genetic variants were selected from genome-wide association studies (GWAS) of smoking behaviour [19, 20]. The variants selected were robustly associated in these studies with smoking heaviness (P<5×10^−8^) and passed UKBB quality control. As far as possible, we used GWAS without data from UKBB to reduce potential statistical bias from winner’s curse [21].

First, we used a single synonymous variant (rs1051730) in the nicotinic receptor gene *CHRNA3*. This had the strongest association with cigarettes smoked per day (CPD) in a study of 73,853 individuals, with follow up in n>140,000 [19] and explains 0.3% of the variance in total smoking amount (log pack years) in UKBB ever smokers. We also used an instrument composed of 4 variants robustly associated with CPD which included the above SNP (Supplement, S2) and explained 0.4% of the variance in smoking heaviness. Finally, we repeated the analysis with 52 SNPs selected from 55 SNPs reported in a recent GWAS with n=337,334 [20] (Supplement, S2), which explained 0.7% of the variance in smoking volume. Data for 120,744/337,344 individuals in that study originated from UKBB.

The single variant rs1051730 and the two genetic instruments have minor and differing associations with confounders; rs1051730 and 4-SNP GRS associate with risky behaviour and the 52-SNP GRS associates with body mass index and socioeconomic position (Supplement, S3). The 52-variant instrument explains the most variance in smoking quantity but also contains UKBB data within the GWAS discovery cohort.

### Analysis Method

We used MR to investigate causality between smoking amount in log pack years and incidence of IPF and COPD. MR relies on several general assumptions [11] which are applied in this case as follows:

➢ the CPD genetic variants are robustly associated with smoking volume as determined by cigarettes per day or log pack years
➢ the CPD genetic variants are not associated, independently of their effects on smoking volume, with confounding factors that bias associations with IPF or COPD
➢ the CPD genetic variants are only associated with IPF or COPD via their effect on smoking volume

Details of the construction of the genetic instruments and statistical method are explained in Supplement S4.

The rs1051730 variant in chromosomal region 15q25 is strongly associated with lung cancer in smokers and several analyses have shown that the association with self-reported smoking heaviness is insufficient to explain that link [22-24]. However, as discussed previously [25], although self-reported smoking heaviness enables the large sample sizes for genetic association studies, it fails to capture smoking exposure fully. We checked that our chosen variants had no direct impact on disease outcomes, by investigating the impact in never smokers.

### Sensitivity Analyses

We carried out a sensitivity study using a COPD definition based on pre-bronchodilator evidence of moderate-to-severe airflow limitation by modified GOLD criteria (FEV1% ≤ 80% predicted and FEV1/FVC ≤ 0.70) [26]. This produced 30,520 unrelated cases, from which we excluded 161 with IPF, leaving 30,359 cases of spirometry-derived COPD for MR analysis.

We also repeated our COPD MR analyses in restricted groups of smoking heaviness (<30 pack years), to address any concerns that COPD sufferers are heavier smokers and the dose-dependent relationship between smoking heaviness and disease prevalence may be non-linear.

## Results

### Demographics

Characteristics of the unrelated 871 IPF cases (cleaned of COPD) and 11,413 COPD cases (cleaned of IPF) are summarised in Table 1. Strong associations when compared with the 366,942 controls were noted between both IPF and COPD and a range of demographic and environmental variables, including older age, male sex, lower socioeconomic position, reduced lung function and reduced exercise. The percentage of IPF ever smokers is lower than reported figures of around 70% [27], possibly because of the female bias in UKBB participants. Strong associations between disease status and ever having smoked were found in both IPF (OR = 1.52 [95%CI:1.32-1.74], P=2.4×10^−8^) and COPD (OR= 5.77 [95%CI:5.48-6.07], P<1×10^−15^), with odds ratios for IPF and COPD in current smokers of 1.66 [95% CI:1.30-2.10], P=3.5×10^−5^ and 15.0 [95%CI:14.1-15.9], P<1×10^−15^ respectively.

**Table 1.**
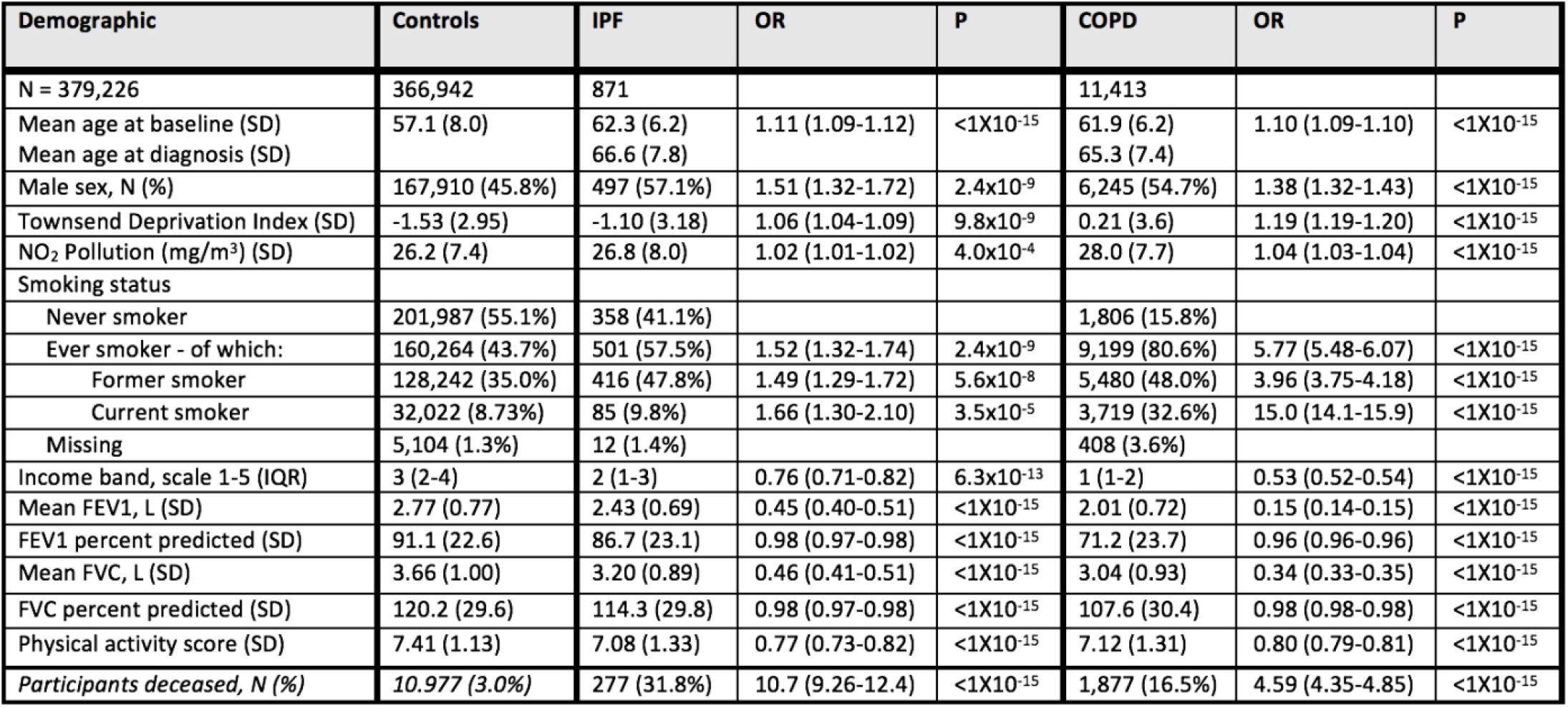
Demographics for IPF and COPD cases compared with controls in UK Biobank, adjusted for age and sex.

### Observational association between smoking heaviness and IPF and COPD

Smoking history in pack years is shown in Figure 2 and Supplement S5. Associations between disease prevalence and smoking heaviness were seen for both conditions; in IPF ever smokers, a unit change in log pack years is linked to a 1.17 higher risk of disease [95%CI:1.05-1.30], p=0.0032 and in COPD ever smokers, a 2.31 higher risk of disease [95%CI:2.26-2.37], p<1×10^−15^. Disease prevalence was associated with smoking heaviness in both current and former smokers, with a larger effect size in COPD (Supplement, S5).

**Figure 2.**
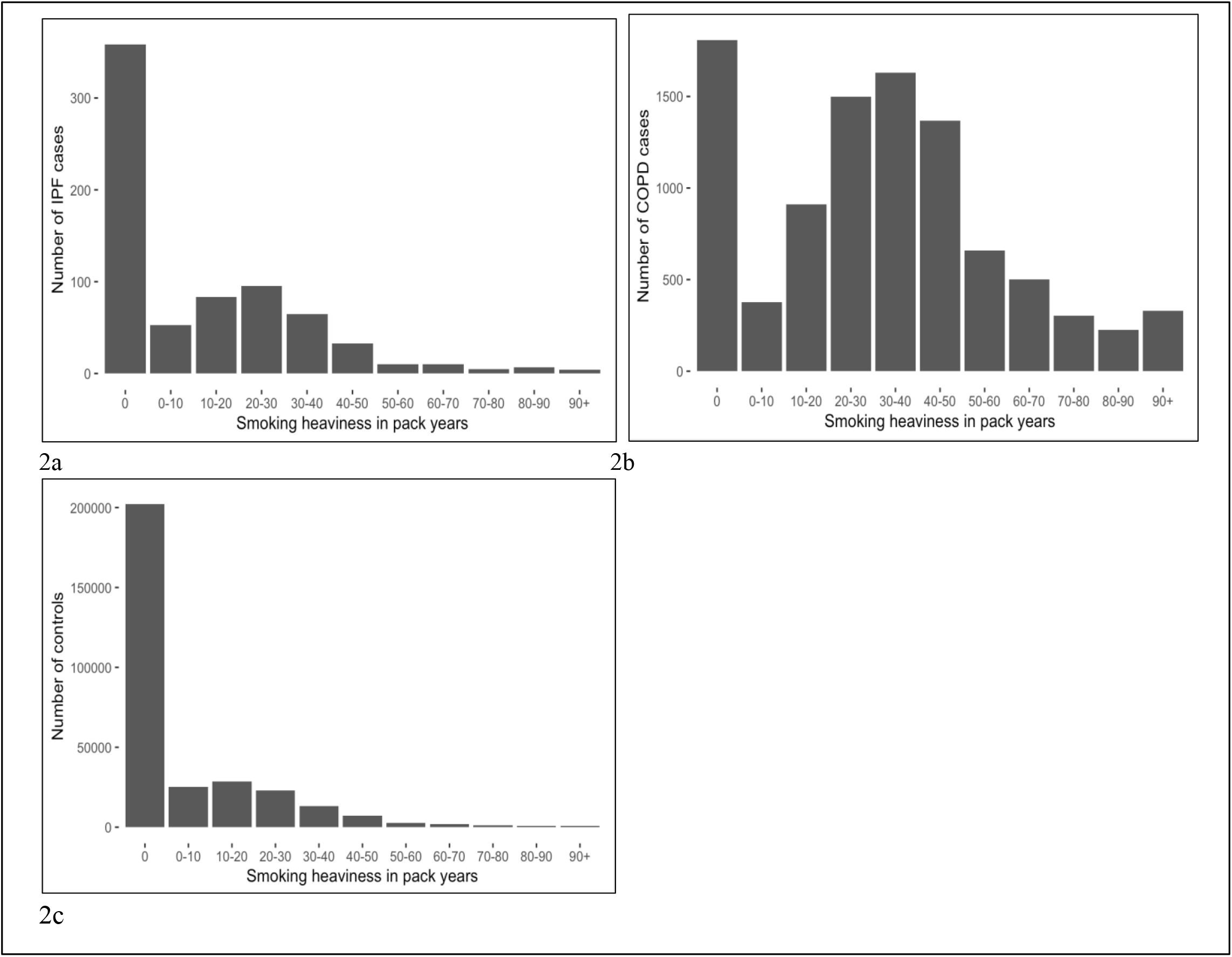
Smoking history in pack years for UK Biobank a) IPF cases, b) COPD cases and c) controls. Number of participants in each smoking band of 10 pack years (pack years = average number of 20-cigarette packs smoked per day X years of smoking), recorded at registration.

Logistic regression showed an association between death and smoking heaviness in log pack years for COPD (OR=1.16; 95%CI:1.08-1.25, p=3.2×10^−5^) but not in IPF (OR=1.04; 95%CI:0.81-1.33, p=0.78), as also reported by the two most recent large observational IPF studies [9, 10]. Despite this, the proportion of participants deceased due to all causes for current/former/never smokers was 20.5%/15.2%/10.6% in COPD and 37.7%/34.1%/27.7% in IPF, showing highest mortality for smokers in both diseases.

### Mendelian randomisation study of smoking causality in IPF and COPD

Using the single smoking variant rs1051730, a genetically instrumented one unit higher smoking volume in log pack years was associated with 7.15 higher odds of COPD in ever smokers, (95%CI:4.73-10.8), P=9.3×10^−21^. Similar results were found in current and former smokers (Supplement, S6). No association was seen between rs1051730 and COPD prevalence amongst never smokers (OR=0.96 [95%CI:0.89-1.03], P=0.24, Supplement, S7). In contrast with COPD, evidence of any causal association in IPF was limited, OR=0.45; 95%CI:0.06-3.28, P=0.43 (Table 2).

**Table 2:**
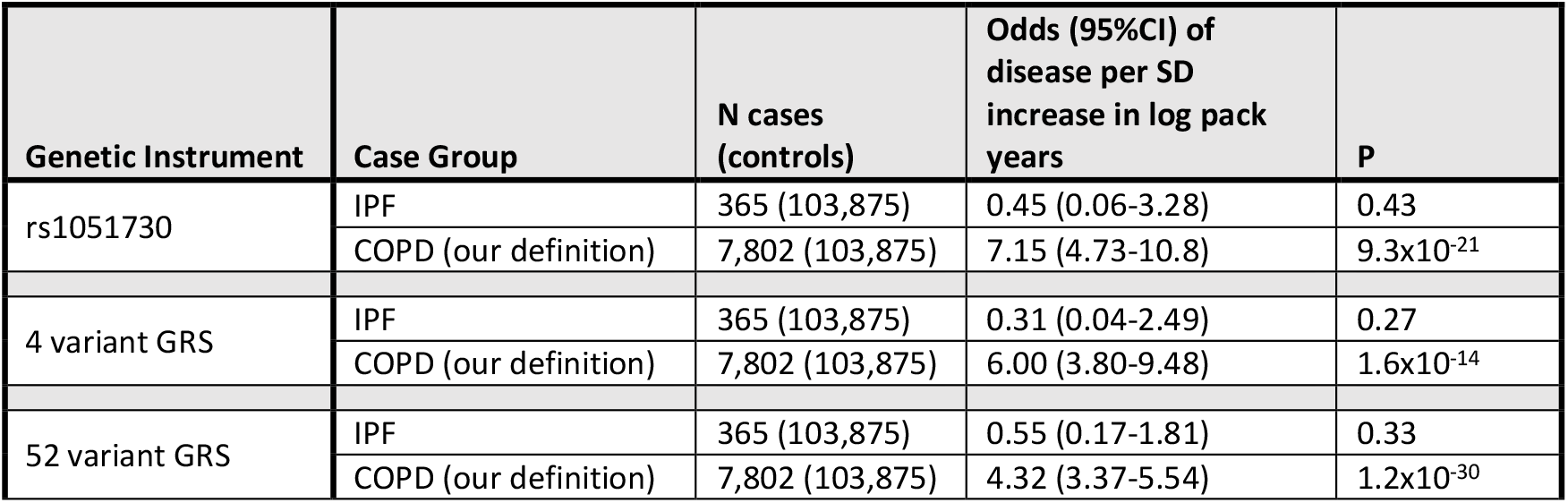
Evidence from UK Biobank data for ever smokers inferring a causal role for smoking in COPD but not in IPF. Associations between smoking heaviness genetic instruments and disease incidence in IPF and COPD for genetic instruments derived from single SNP, 4 variants and 52 variants as detailed in the supplement. Results were adjusted for age, sex, ancestral principal components, assessment centre and genotyping platform. The number of cases and controls is reduced for the MR analyses due to missing smoking volume data. Further results and spirometry-defined COPD are included in the supplement.

Disease prevalence associates with number of rs1051730 alleles in COPD in former and current smokers, but not in never smokers. No association was evident across smoking strata for IPF (Figure 3).

**Figure 3.**
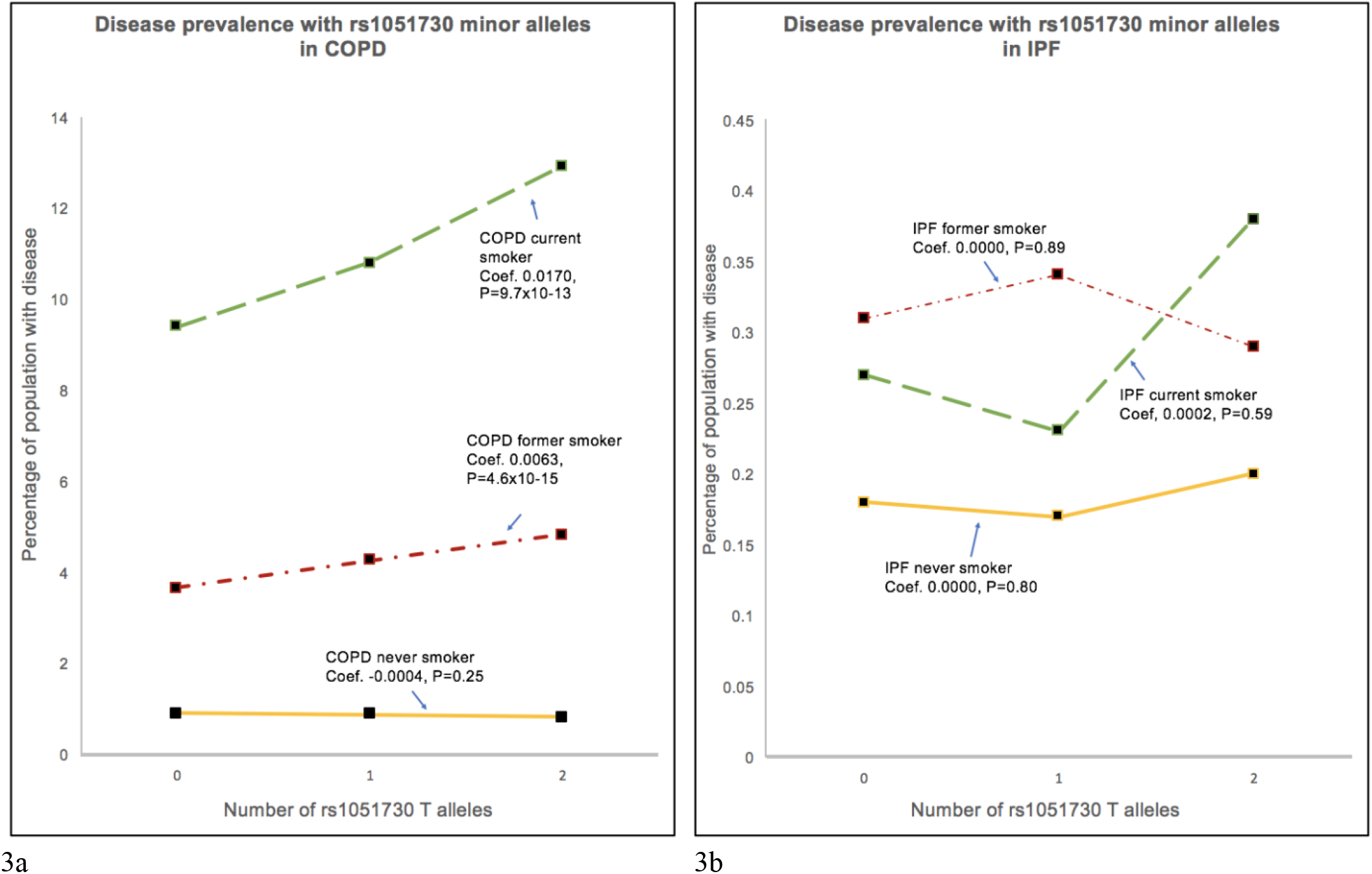
Dependence of disease prevalence for a) COPD and b) IPF on number of smoking linked SNP rs1051730 risk alleles for current, former and never smokers (note different scales). In COPD, disease incidence increases with the number of copies of the smoking volume risk allele (T). This is most pronounced in current smokers, less pronounced in former smokers and not evident in non-smokers. In IPF, disease incidence is not associated with the number of rs1051730 C/T risk alleles affecting volume of smoking. Regression coefficients and P values are shown in each case.

Using the 4-variant genetic risk score, a unit increase in smoking volume GRS was associated with higher odds of COPD in ever smokers, OR=6.00 [95%CI:3.80-9.48], P=1.6×10^−14^. Again, no association was seen in IPF (Table 2, Figure 4). Using the 52-variant CPD genetic risk score, a unit increase in smoking volume GRS was associated with higher odds of COPD in ever smokers, OR=4.32; 95%CI:3.37-5.54, P=1.2×10^−30^, with no association seen in IPF (Table 2, Figure 4).

**Figure 4.**
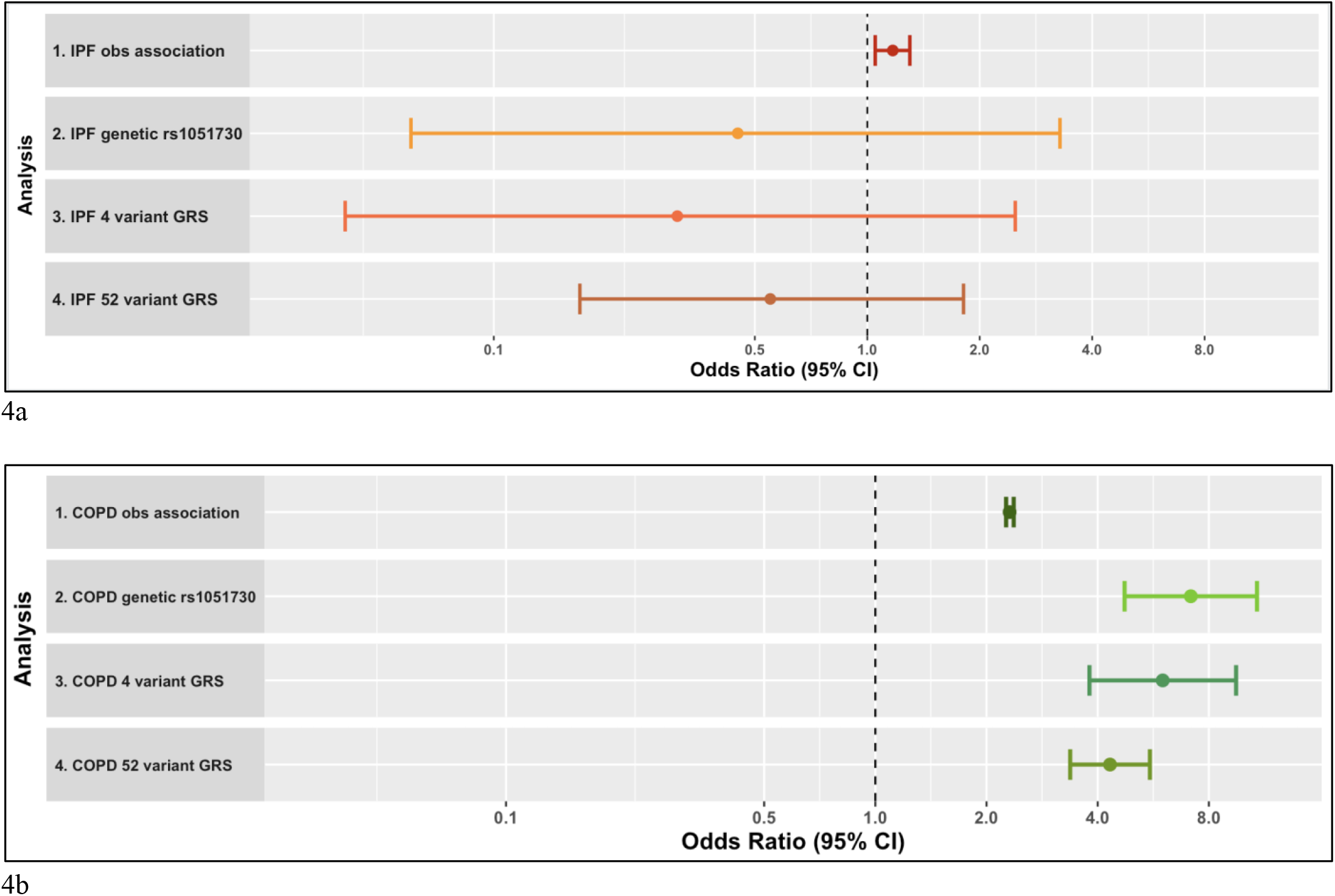
Observational associations and genetically derived causal associations for smoking heaviness amongst ever smokers in IPF and COPD. (a) Associations with smoking heaviness for IPF: 1. Observational association of disease with log pack years, 2-4. Genetically derived causal association of disease with log pack years using genetic variant rs1051730, 4-variant genetic risk score, 52-variant genetic risk score respectively (b) Associations with smoking heaviness for COPD: 1. Observational association of disease with log pack years, 2-4. Genetically derived causal association of disease with log pack years using genetic variant rs1051730, 4-variant genetic risk score, 52-variant genetic risk score respectively.

Our chosen variants had no direct impact on disease outcomes (Supplement S7; associations in never and ever smokers). Sensitivity analysis, with COPD defined using lung function demonstrated similar results (Supplement, S6). Results for the MR analysis for COPD at lower smoking heaviness (<30 pack years) also remained significant with higher odds ratios (Supplement, S8).

### Observational associations between parental smoking and IPF

In the full dataset, ever smoking was associated with the only direct data for parental smoking, namely ‘mother smoking during pregnancy’ (OR=1.09; 95%CI:1.08-1.11, p<10^−15^). Ever smoking was also associated with paternal lung cancer prevalence as a possible indicator for father smoking (OR=1.08; 95%CI:1.05-1.10, p=1.9×10^−9^). Paternal lung cancer associates with maternal smoking during pregnancy (OR=1.48; 95%CI:1.44-1.52, p<10^−15^), suggesting that the father is also likely to have a history of smoking (active or passive), if the mother smoked during pregnancy.

IPF prevalence was also associated with maternal smoking during pregnancy (OR=1.31; 95%CI:1.12-1.52, p=5.9×10^−4^). We therefore see evidence of an association between parental smoking and IPF in the limited data that are available. Due to small numbers in the IPF cohort, (31/782 for maternal lung cancer and 80/775 for paternal lung cancer), we had insufficient power to deduce whether analyses of parental lung cancer prevalence showed an association with IPF.

## Discussion

Associations between disease prevalence and smoking were observed in UKBB for both IPF and COPD compared with controls. Mendelian randomisation provided evidence inferring a causal link between smoking and COPD but there was limited evidence of any causal link between smoking and IPF. COPD disease prevalence amongst smokers was dependent on the number of variant alleles of the SNP linked with smoking volume, rs1051730, but this was not the case in IPF. These findings provide further evidence of smoking causality in COPD but little evidence of causality in IPF.

A previous meta-analysis reported that variation in the rs1051730 A allele used in two of our MR analyses, was associated with an increased risk of COPD regardless of smoking exposure [28], raising the question of whether the variant may play a role in COPD pathogenesis alongside smoking. In that review, for groups with similar reported smoking exposure (∼40 pack years), disease prevalence was higher in subjects with a variant allele. It was suggested that the A allele predisposes smokers to COPD, possibly via the inflammatory response, and that the dose relationship with smoking may not be linear. A recent MR study into systemic inflammation with smoking reported that the rs1051730 SNP was not associated with most potential confounders whereas tobacco smoking was associated with almost all and concluded that the association with lung function measurements should be interpreted as downstream effects of tobacco smoking rather than potential confounders [13]. Furthermore, studies of this SNP have reported a disconnect between self-reported smoking quantities and concentrations of nicotine metabolites [29]. In addition to using different genetic instruments (with similar results), we have further addressed these concerns with our sensitivity study at smoking heaviness <30 pack years for COPD, similar to IPF. We also showed that no disease dependence of this risk allele was found in non-smokers with COPD, supporting previous evidence that the variant has no causal effect other than through smoking [30].

A limitation of our study, as with many IPF studies, is the relatively small number of IPF cases (especially when cleaned of COPD), leading to underpowered MR analyses. Power calculations for MR analyses [31] showed low values for IPF as a result of the low observational odds ratio of 1.17 (with F statistics of 241-873). If the observed effect size had been similar to that found for COPD (2.31), the power values would have almost sufficed (0.62, where ≥0.8 is ideal); with the observed OR of 1.17, it would require around 36000 IPF cases to achieve satisfactory power. However, since the odds ratio values for the causal effect of smoking in IPF were consistently less than 1 across different analyses, this provides a measure of confidence that increased numbers would not produce statistically significant odds ratios greater than 1, as was seen for COPD. Our study is also limited by the MR assumptions described earlier. We have mitigated the issue of confounding with the use of three instruments and additional sensitivity studies. Although there was some potential pleiotropy, with for example risky behaviour, this differed between the three instruments so the consistent findings strengthen the assumption that the effect found in COPD is not due to pleiotropy. Additional limitations include that the UKBB data set has a healthy volunteer bias and smoking data is gathered at recruitment rather than at diagnosis. For these reasons, we recommend further analysis using 2-sample MR methods in the future, if more data become available.

Subject to these limitations and cognisant of the fact that absence of evidence is not evidence of absence, we have presented limited evidence suggesting that, despite a well-established association with disease incidence (replicated here), smoking may not cause IPF. This suggestion could be an important one for the field of fibrosing lung diseases, in which smoking is a well-established risk factor, and there are two important consequences. First, the overriding message remains that smokers diagnosed with IPF should be encouraged to stop smoking due to the higher all cause mortality amongst smokers. In COPD the case is even stronger, with several reports [32], of higher mortality in current smokers. Second, if smoking is not causal for IPF, we need to consider correlation without causality and there are many possible explanations. One might be that smoking initiation triggers a response to be ‘primed’ and beyond that, neither increased smoking volume (to which these genetic instruments link) nor smoking cessation have an impact on disease incidence or progression. Alternatively, there may be other confounding factors in our analyses, such as workplace inhalational hazards that are more common amongst smokers. A recent review concluded that workplace exposures contribute to 26% of IPF cases [33]. Nor can we exclude any impacts of passive smoking.

Another more complex hypothesis is one of inherited response to the threat of smoking (and lung cancer in particular), transmitted with greatest efficacy through the paternal line. There is evidence in other fields, of multigenerational epigenetic adaptation to similar environmental cues, such as inherited hepatic regulation of wound-healing in response to ancestral alcohol-induced liver fibrosis [34]. Adolescent and parental smoking populations are linked [35], with offspring of smoking parents more likely to smoke, as supported by our findings and also supported by an observed association for IPF with maternal smoking (with data only available for smoking during pregnancy). It is therefore possible that in some cases, IPF may be caused by epigenetic changes inherited as a result of ancestral smoking to confer cancer protection. To some extent, this aligns with George Williams’ concept of aging and senescence caused by ‘antagonistic pleiotropy’, where induced changes that are protective in early life have negative consequences after the reproductive years [36]. This would explain why smoking cessation appears to have little impact on disease incidence or progression.

In summary, we have shown evidence inferring that smoking is causal for COPD but a lack of corresponding evidence inferring that smoking is causal for IPF.

## Data Availability

All data used in analyses are available upon request from UK Biobank at https://www.ukbiobank.ac.uk

## Acknowledgements and funding

A. Duckworth is funded by the GW4 MRC Doctoral Training Partnership.

M. Gibbons has received support to attend conferences and professional fees from Roche and Boehringer-Ingelheim.

J. Tyrrell is supported by an Academy of Medical Sciences (AMS) Springboard award, which is supported by the AMS, the Wellcome Trust, GCRF, the Government Department of Business, Energy and Industrial strategy, the British Heart Foundation and Diabetes UK [SBF004\1079].

This research has been conducted using the UK Biobank Resource (applications 9072 and 44046).

