## Supplementary Material for "A Mendelian randomisation study of smoking causality in IPF compared with COPD"

[1] Institute of Biomedical & Clinical Science, College of Medicine & Health, University of Exeter, Exeter, UK
[2] Exeter Patients in Collaboration (EPIC) for PF, Exeter, UK
[3] Respiratory Medicine Department, Royal Devon and Exeter NHS Foundation Trust, Exeter, UK
[4] Department of Pharmacy and Pharmacology, University of Bath, Bath, UK
[5] These authors contributed equally

| Demographic | Controls | IPF | OR | P | COPD | OR | P |
|---|---|---|---|---|---|---|---|
| N = 379,226 | 366,942 | 871 | | | 11,413 | | |
| Mean age at baseline (SD)<br>Mean age at diagnosis (SD) | 57.1 (8.0) | 62.3 (6.2)<br>66.6 (7.8) | 1.11 (1.09-1.12) | <1X10$^{-15}$ | 61.9 (6.2)<br>65.3 (7.4) | 1.10 (1.09-1.10) | <1X10$^{-15}$ |
| Male sex, N (%) | 167,910 (45.8%) | 497 (57.1%) | 1.51 (1.32-1.72) | 2.4x10$^{-9}$ | 6,245 (54.7%) | 1.38 (1.32-1.43) | <1X10$^{-15}$ |
| Townsend Deprivation Index (SD) | -1.53 (2.95) | -1.10 (3.18) | 1.06 (1.04-1.09) | 9.8x10$^{-9}$ | 0.21 (3.6) | 1.19 (1.19-1.20) | <1X10$^{-15}$ |
| $NO_2$ Pollution (mg/m$^3$) (SD) | 26.2 (7.4) | 26.8 (8.0) | 1.02 (1.01-1.02) | 4.0x10$^{-4}$ | 28.0 (7.7) | 1.04 (1.03-1.04) | <1X10$^{-15}$ |
| Smoking status | | | | | | | |
| Never smoker | 201,987 (55.1%) | 358 (41.1%) | | | 1,806 (15.8%) | | |
| Ever smoker - of which: | 160,264 (43.7%) | 501 (57.5%) | 1.52 (1.32-1.74) | 2.4x10$^{-9}$ | 9,199 (80.6%) | 5.77 (5.48-6.07) | <1X10$^{-15}$ |
| Former smoker | 128,242 (35.0%) | 416 (47.8%) | 1.49 (1.29-1.72) | 5.6x10$^{-8}$ | 5,480 (48.0%) | 3.96 (3.75-4.18) | <1X10$^{-15}$ |
| Current smoker | 32,022 (8.73%) | 85 (9.8%) | 1.66 (1.30-2.10) | 3.5x10$^{-5}$ | 3,719 (32.6%) | 15.0 (14.1-15.9) | <1X10$^{-15}$ |
| Missing | 5,104 (1.3%) | 12 (1.4%) | | | 408 (3.6%) | | |
| Income band, scale 1-5 (IQR) | 3 (2-4) | 2 (1-3) | 0.76 (0.71-0.82) | 6.3x10$^{-13}$ | 1 (1-2) | 0.53 (0.52-0.54) | <1X10$^{-15}$ |
| Mean FEV1, L (SD) | 2.77 (0.77) | 2.43 (0.69) | 0.45 (0.40-0.51) | <1X10$^{-15}$ | 2.01 (0.72) | 0.15 (0.14-0.15) | <1X10$^{-15}$ |
| FEV1 percent predicted (SD) | 91.1 (22.6) | 86.7 (23.1) | 0.98 (0.97-0.98) | <1X10$^{-15}$ | 71.2 (23.7) | 0.96 (0.96-0.96) | <1X10$^{-15}$ |
| Mean FVC, L (SD) | 3.66 (1.00) | 3.20 (0.89) | 0.46 (0.41-0.51) | <1X10$^{-15}$ | 3.04 (0.93) | 0.34 (0.33-0.35) | <1X10$^{-15}$ |
| FVC percent predicted (SD) | 120.2 (29.6) | 114.3 (29.8) | 0.98 (0.97-0.98) | <1X10$^{-15}$ | 107.6 (30.4) | 0.98 (0.98-0.98) | <1X10$^{-15}$ |
| Physical activity score (SD) | 7.41 (1.13) | 7.08 (1.33) | 0.77 (0.73-0.82) | <1X10$^{-15}$ | 7.12 (1.31) | 0.80 (0.79-0.81) | <1X10$^{-15}$ |
| *Participants deceased, N (%)* | *10.977 (3.0%)* | 277 (31.8%) | 10.7 (9.26-12.4) | <1X10$^{-15}$ | 1,877 (16.5%) | 4.59 (4.35-4.85) | <1X10$^{-15}$ |

**Table 1 Demographics for IPF and COPD cases compared with controls in UK Biobank, adjusted for age and sex**

**Observational association between smoking heaviness and IPF and COPD**

Smoking history in pack years is shown in Figure 2 and Supplement S5. Associations between disease prevalence and smoking heaviness were seen for both conditions; in IPF ever smokers, a unit change in log pack years is linked to a 1.17 higher risk of disease [95%CI:1.05-1.30], p=0.0032 and in COPD ever smokers, a 2.31 higher risk of disease [95%CI:2.26-2.37], p<1x10$^{-15}$. Disease prevalence was associated with smoking heaviness in both current and former smokers, with a larger effect size in COPD (Supplement, S5).

Logistic regression showed an association between death and smoking heaviness in log pack years for COPD (OR=1.16; 95%CI:1.08-1.25, p=3.2x10$^{-5}$) but not in IPF (OR=1.04; 95%CI:0.81-1.33, p=0.78), as also reported by the two most recent large observational IPF studies [9, 10]. Despite this, the proportion of participants deceased due to all causes for current/former/never smokers was 20.5%/15.2%/10.6% in COPD and 37.7%/34.1%/27.7% in IPF, showing highest mortality for smokers in both diseases.

**Mendelian randomisation study of smoking causality in IPF and COPD**
Using the single smoking variant rs1051730, a genetically instrumented one unit higher smoking volume in log pack years was associated with 7.15 higher odds of COPD in ever smokers, (95%CI:4.73-10.8), P=9.3x10$^{-21}$. Similar results were found in current and former smokers (Supplement, S6). No association was seen between rs1051730 and COPD prevalence amongst never smokers (OR=0.96 [95%CI:0.89-1.03], P=0.24, Supplement, S7). In contrast with COPD, evidence of any causal association in IPF was limited, OR=0.45; 95%CI:0.06-3.28, P=0.43 (Table 2).

| Genetic Instrument | Case Group | N cases (controls) | Odds (95%CI) of disease per SD increase in log pack years | P |
|---|---|---|---|---|
| rs1051730 | IPF | 365 (103,875) | 0.45 (0.06-3.28) | 0.43 |
| | COPD (our definition) | 7,802 (103,875) | 7.15 (4.73-10.8) | 9.3x10$^{-21}$ |
| 4 variant GRS | IPF | 365 (103,875) | 0.31 (0.04-2.49) | 0.27 |
| | COPD (our definition) | 7,802 (103,875) | 6.00 (3.80-9.48) | 1.6x10$^{-14}$ |
| 52 variant GRS | IPF | 365 (103,875) | 0.55 (0.17-1.81) | 0.33 |
| | COPD (our definition) | 7,802 (103,875) | 4.32 (3.37-5.54) | 1.2x10$^{-30}$ |

**Table 2: Evidence from UK Biobank data for ever smokers inferring a causal role for smoking in COPD but not in IPF.** Associations between smoking heaviness genetic instruments and disease incidence in IPF and COPD for genetic instruments derived from single SNP, 4 variants and 52 variants as detailed in the supplement. Results were adjusted for age, sex, ancestral principal components, assessment centre and genotyping platform. The number of cases and controls is reduced for the MR analyses due to missing smoking volume data. Further results and spirometry-defined COPD are included in the supplement.

This research has been conducted using the UK Biobank Resource (applications 9072 and 44046).

**Figure 1 Principle of Mendelian Randomisation** (a) Illustration demonstrating how Mendelian inheritance leads to a random allocation of variant alleles amongst offspring such that confounders affecting different social groups across the whole sample can be controlled for statistically; akin to a natural randomised control trial (b) If a trait such as smoking intensity (X) plays a causal role in the disease outcome IPF or COPD (Y), genetic variants ($Z_i$) associated with smoking intensity will also be associated with the outcome Y. Since genotype is assigned at conception, it should not be associated with environmental risk factors that would normally confound the association between the trait and lung disease (eg diet or pollution). Weighted estimates of the genetic-trait association (w) and the genetic-lung disease association (x) can be used to infer the causal effect of that trait on the lung disease (y=x/w), which is expected to be free from confounding.

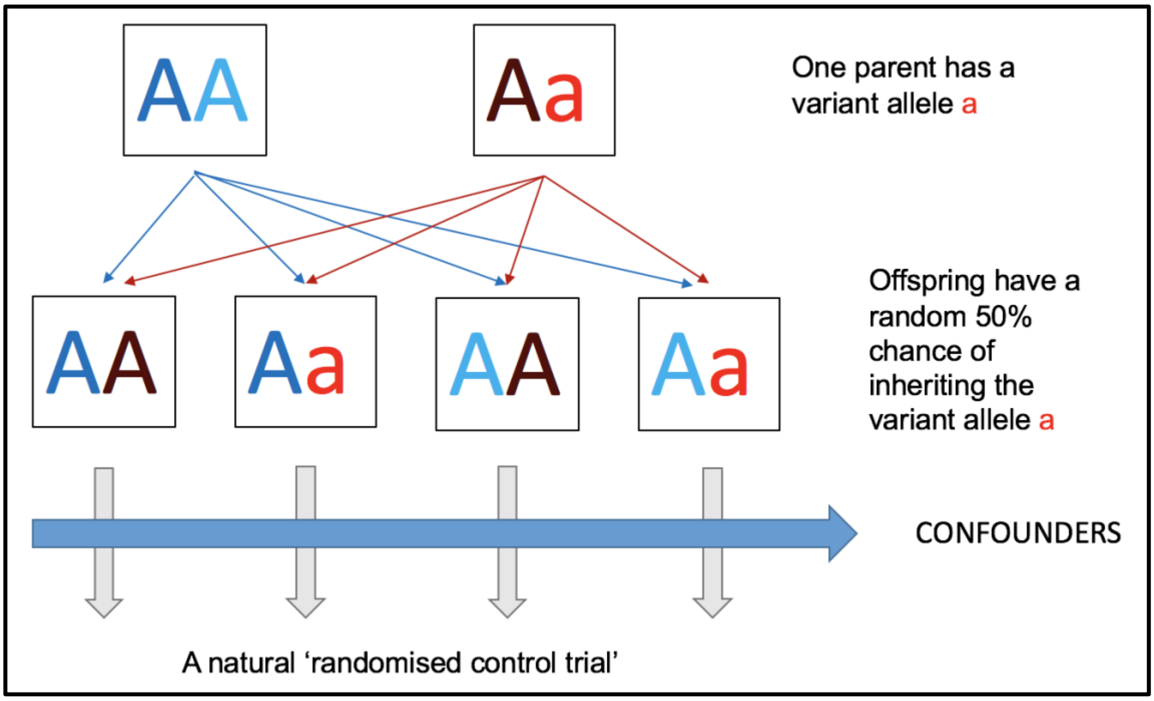

1a

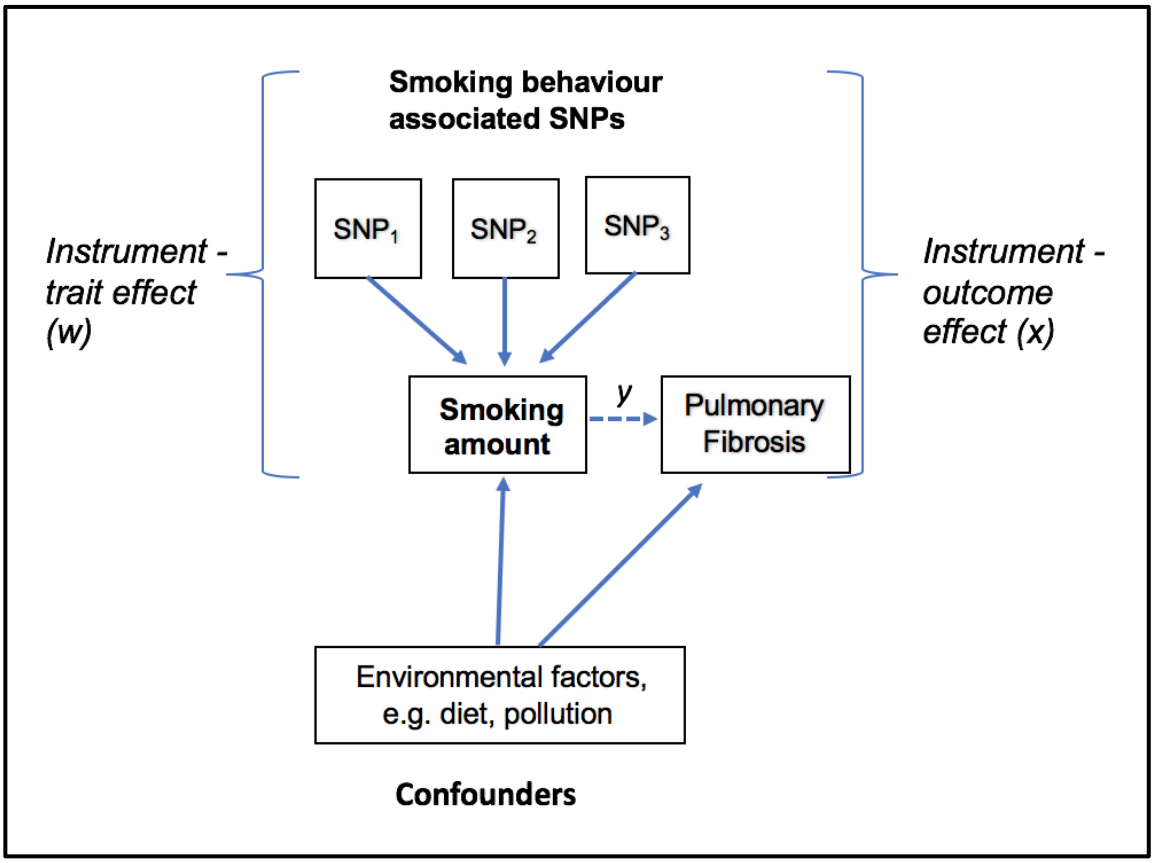

1b

**Figure 2 Smoking history in pack years for UK Biobank a) IPF cases, b) COPD cases and c) controls.** Number of participants in each smoking band of 10 pack years (pack years = average number of 20-cigarette packs smoked per day X years of smoking), recorded at registration.

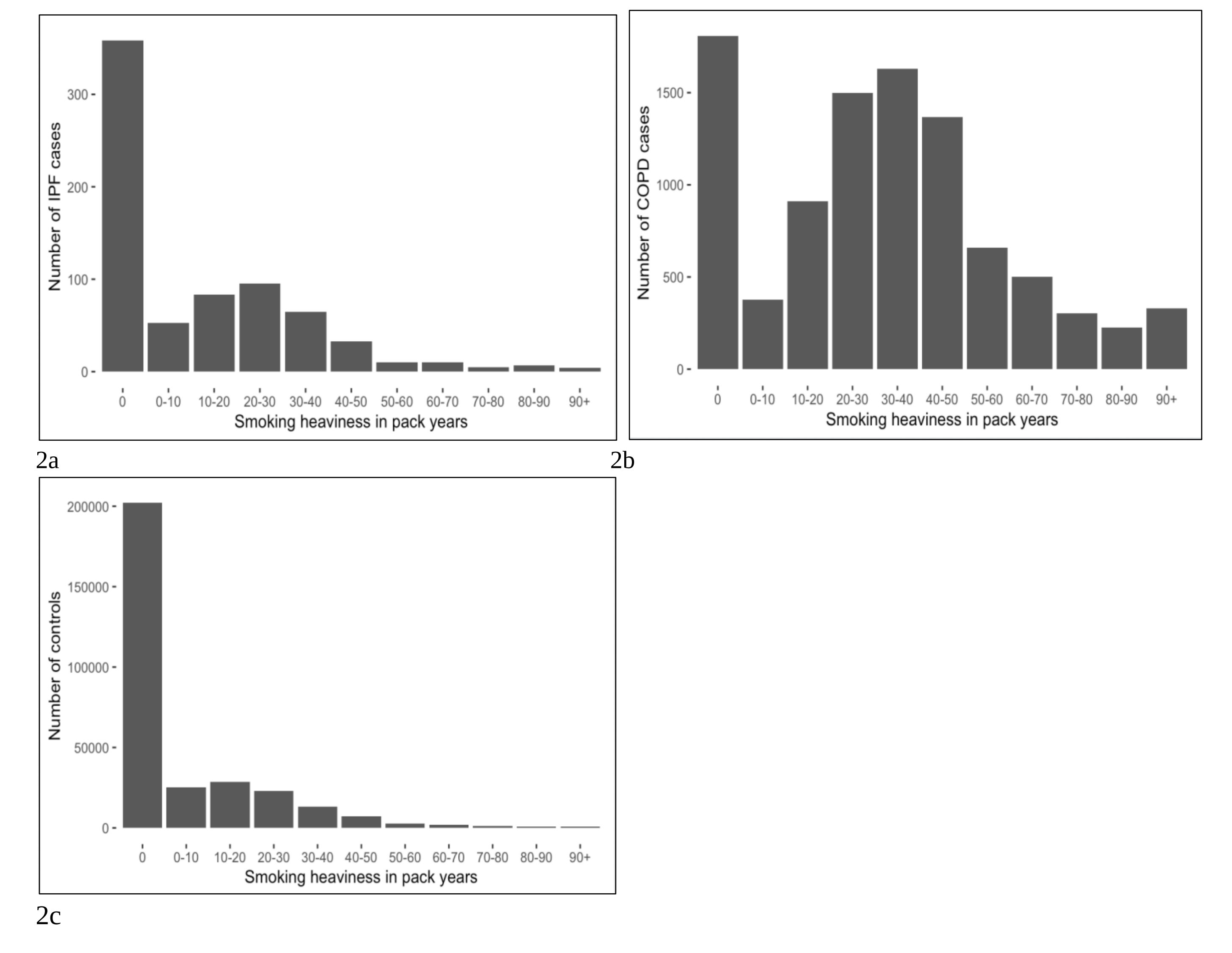

**Figure 3 Dependence of disease prevalence for a) COPD and b) IPF on number of smoking linked SNP rs1051730 risk alleles for current, former and never smokers (note different scales).** In COPD, disease incidence increases with the number of copies of the smoking volume risk allele (T). This is most pronounced in current smokers, less pronounced in former smokers and not evident in non-smokers. In IPF, disease incidence is not associated with the number of rs1051730 C/T risk alleles affecting volume of smoking. Regression coefficients and P values are shown in each case.

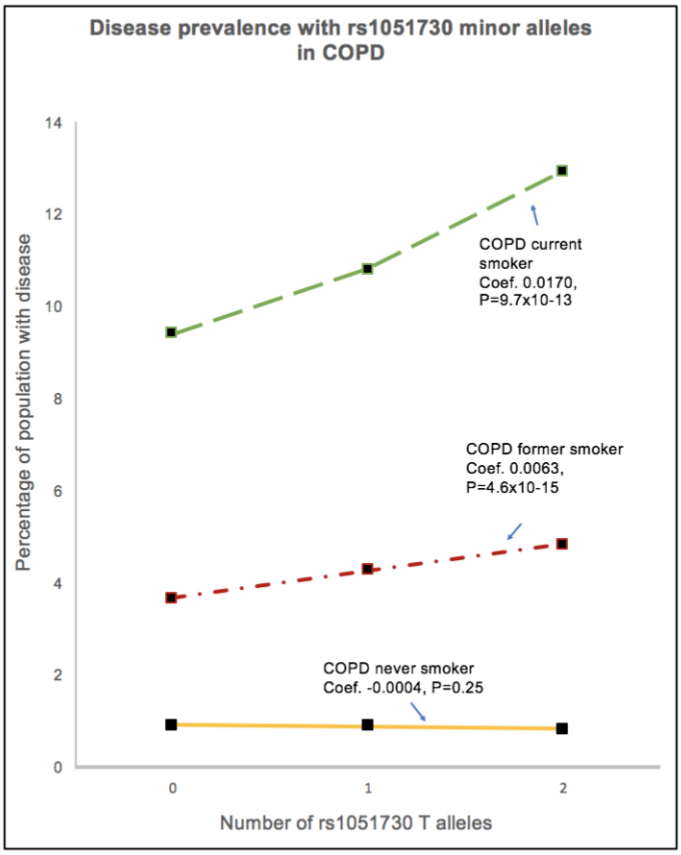

3a

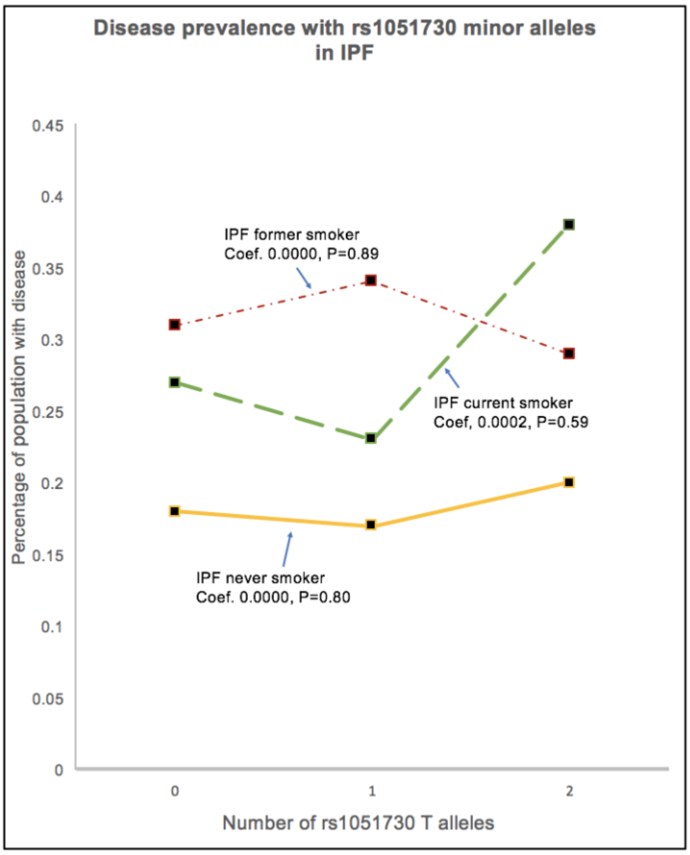

3b

**Figure 4. Observational associations and genetically derived causal associations for smoking heaviness amongst ever smokers in IPF and COPD (**a) Associations with smoking heaviness for IPF: 1. Observational association of disease with log pack years, 2-4. Genetically derived causal association of disease with log pack years using genetic variant rs1051730, 4-variant genetic risk score, 52-variant genetic risk score respectively (b) Associations with smoking heaviness for COPD: 1. Observational association of disease with log pack years, 2-4. Genetically derived causal association of disease with log pack years using genetic variant rs1051730,  4-variant genetic risk score, 52-variant genetic risk score respectively.

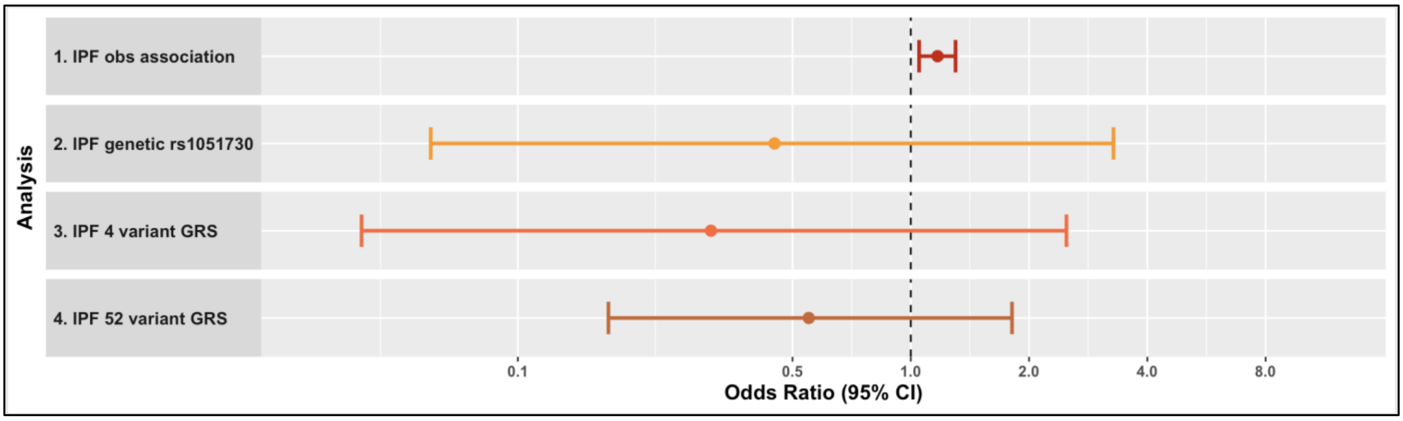

4a

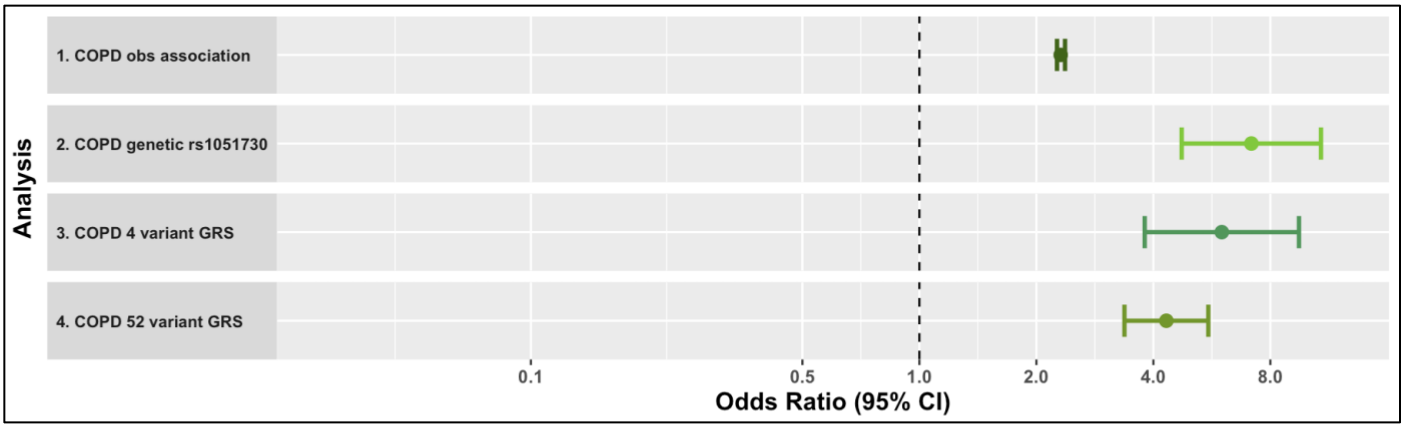

4b
